# A single mRNA immunization boosts cross-variant neutralizing antibodies elicited by SARS-CoV-2 infection

**DOI:** 10.1101/2021.02.05.21251182

**Authors:** Leonidas Stamatatos, Julie Czartoski, Yu-Hsin Wan, Leah J. Homad, Vanessa Rubin, Hayley Glantz, Moni Neradilek, Emilie Seydoux, Madeleine F. Jennewein, Anna J. MacCamy, Junli Feng, Gregory Mize, Stephen C. De Rosa, Andrés Finzi, Maria P. Lemos, Kristen W. Cohen, Zoe Moodie, M. Juliana McElrath, Andrew T. McGuire

## Abstract

Emerging SARS-CoV-2 variants have raised concerns about resistance to neutralizing antibodies elicited by previous infection or vaccination. We examined whether sera from recovered and naïve donors collected prior to, and following immunizations with existing mRNA vaccines, could neutralize the Wuhan-Hu-1 and B.1.351 variants. Pre-vaccination sera from recovered donors neutralized Wuhan-Hu-1 and sporadically neutralized B.1.351, but a single immunization boosted neutralizing titers against all variants and SARS-CoV-1 by up to 1000-fold. Neutralization was due to antibodies targeting the receptor binding domain and was not boosted by a second immunization. Immunization of naïve donors also elicited cross-neutralizing responses, but at lower titers. Our study highlights the importance of vaccinating both uninfected and previously infected persons to elicit cross-variant neutralizing antibodies.

## MAIN

The SARS-CoV-2 betacoronavirus first emerged in the Hubei Province of China in late 2019 and has since infected over 115 million people and caused over 2.5 million deaths in 192 countries (*1-3*).

Infection is mediated by the viral spike protein (S) which is comprised of an S1 domain that contains a N-terminal domain (NTD), a C-terminal domain (CTD), and a receptor binding domain (RBD) which mediates attachment to the entry receptor angiotensin converting enzyme 2 (ACE2), and an S2 domain that contains the fusion machinery (*4-8*).

Pre-existing immunity to SARS-CoV-2 is associated with protection against re-infection in humans (*9-11*) and in non-human primates (*12, 13*). Although the correlates of protection in humans against repeat infection or following vaccination have not been firmly established, neutralizing antibodies (nAbs) are thought to be an important component of a protective immune response against SARS-CoV-2 (*14, 15*). In support of this, passive transfer of nAbs limits respiratory tract infection and protects against infection in animal models (*16-20*) and may contribute to protection against infection in humans (*9*). SARS-CoV-2 infection rapidly elicits nAbs (*16, 21-24*) that decline, but remain detectable over several months (*25-29*).

The majority of serum neutralizing antibody responses elicited during natural infection are directed at the receptor bidning domain (RBD) (*21, 23, 30, 31*). Numerous neutralizing anti-RBD monoclonal antibodies (mAbs) have been characterized, the most potent of which block the RBD-ACE2 interaction (*16, 17, 22-24, 32-37*). Neutralizing against other region of the viral spike have also been identified (*24, 33, 38-42*).

Two mRNA-based vaccines (Pfizer/BioNTech BNT162b2, and Moderna mRNA-1273) have received emergency use authorization in several countries. Both encode a stabilized ectodomain version of the S protein derived from the Wuhan-Hu-1 variant isolated in December 2019 (*43*), show greater than 94% efficacy at preventing COVID-19 illness (*44-47*), and elicit nAbs (*48, 49*).

Due to the high global burden of SARS-CoV-2 transmission, viral evolution is occurring. Recently, viral variants of concern have emerged in the United Kingdom (B.1.1.7), South Africa (B.1.351), and Brazil (P.1) that harbor specific mutations in their S proteins that may be associated with increased transmissibility (*50-55*).

Of particular concern are mutations found in the B.1.351 lineage, which is defined by the D80A and D215G in the N-terminal domain (NTD), and the K417N, E484K, N501Y mutations in the RBD and the D614G mutation in S2 (*52, 56*). An A701V mutation in S2 is also observed at high frequencies, while deletions in 242-244 and a R246I mutation in the NTD and a mutation in the leader peptide (L18F) are present at lower frequencies (*52*).

The B.1.1.7, B.1.351, and P.1 lineages all harbor a N501Y mutation in the RBD which increases the affinity for the ACE2 receptor (*57, 58*), and a D614G mutation which increases virion spike density, infectivity and transmissibility (*59, 60*). The B.1.351 and P.1 lineages also share the E484K mutations in the RBD and both variants are mutated at 417 (K417T in P.1).

Mutations found in emergent S variants decrease sensitivity to neutralization by mAbs, convalescent plasma, and sera from vaccinated individuals (*27, 37, 58, 61-70*). As a result, there is concern that these and other emerging variants can evade neutralizing antibody responses generated during infection with variants circulating earlier in the pandemic and also from neutralizing antibody responses elicited by vaccines based on the spike protein of the Wuhan-Hu-1 variant. Indeed, there is concern that these mutations are responsible for reduced efficacy observed in ongoing trials of SARS-CoV-2 vaccines in South Africa (*71, 72*).

Here, we evaluated the neutralization susceptibility of spike variants harboring lineage-defining and prevalent B.1.351 mutations to sera from 15 donors with previously confirmed SARS-CoV-2 infection (herein referred to as previously infected donors or PIDs), that were collected prior to, and following one or two immunizations with either mRNA vaccine, or from 13 uninfected donors who received two doses of the above vaccines (herein called naïve donors or NDs; Tables S1 and S2), as well as anti-spike mAbs isolated from infected but not vaccinated patients.

Antibody neutralization experiments were performed with pseudoviruses expressing either the full-length Wuhan-Hu-1 S, or either of two versions of the B.1.351 lineage S, one herein referred to as B.1.351 containing the lineage-defining S mutations D80A, D215G, K417N, E484K, N501Y and D614G mutations and the A701V mutation that is highly prevalent in this lineage, and a second variant that also includes a Δ242-243 deletion (B.1.351Δ242-243). The viral stocks were appropriately diluted to achieve comparable entry levels during the neutralization experiments (Fig. S1).

We first evaluated the neutralizing potency of several mAbs isolated from non-vaccinated patients infected early in the pandemic, which target different epitopes: three against the RBD (CV30, CV3-1 and CV2-75) and one against the NTD (CV1) (Fig. S2). CV30 is a member of the VH3-53 class of antibodies that bind to the receptor binding motif (RBM) (*22, 32, 73-78*). It makes direct contact with the K417 and N501 residues in the RBM that are mutated in the B.1.351 and P.1 lineages, but unlike other known VH3-53 mAbs it does not contact E484 (*78*).

The neutralization potency of this mAb was ∼10-fold weaker towards both B.1.351 variants (Fig. 1A). Similarly, the non-VH3-53 mAb CV3-1 was 3-4-fold less potent against the B.1.351 variants (Fig. 1B), while CV2-75 was modestly less effective (Fig. 1C). In contrast, the anti-NTD CV1 mAb was unable to neutralize either B.1.351 variant (Fig. 1D). As expected, the control anti-EBV mAb AMMO1 was non-neutralizing (*79*) (Fig. 1E). Collectively, these data indicate that the B.1.351 variants tested here are more resistant to neutralization by mAbs isolated from subjects infected by viral variants from early in the pandemic. We therefore examined whether the B.1.351 variants are resistant to neutralizing antibody responses elicited by the Pfizer/BioNTech or Moderna mRNA vaccines, in PIDs and NDs.

**Figure 1.**
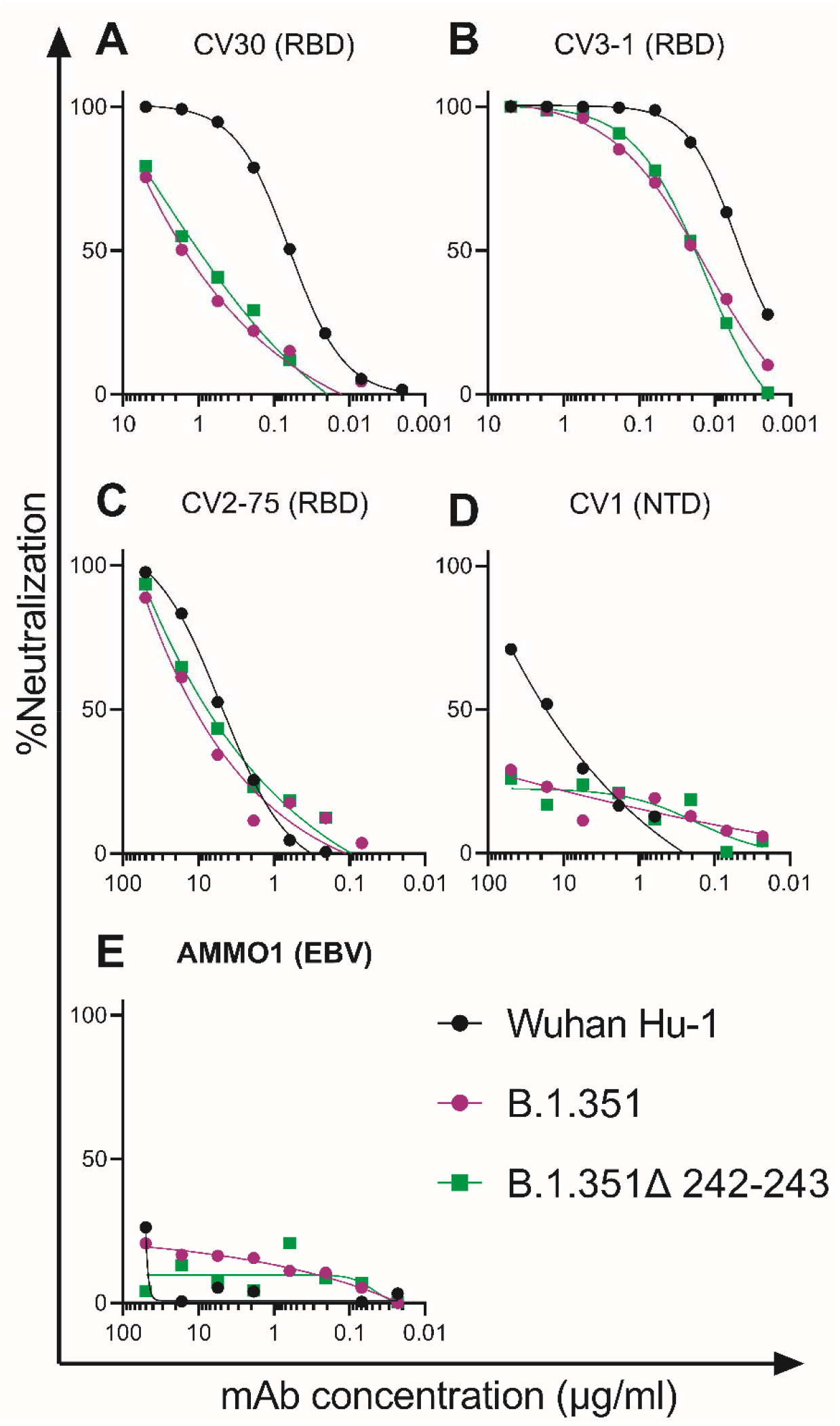
B.1.351 variants show decreased susceptibility to neutralizing monoclonal anibodies. The ability of the indicated monoclonal antibodies (mAbs) to neutralize Wuhan-Hu-1, B.1.351 and B.1.351Δ242-243 pseudovirus infectivity in 293T-hACE2 cells was measured as indicated. The epitope specificity of each mAb is shown in parentheses (RBD: receptor binding domain; NTD: N-terminal domain; EBV: Epstein-Barr virus). Data points represent the mean of two techincal replicates. Data is represenative of two independent experiments.

The RBD-specific IgG, IgM and IgA binding responses to the RBD from the Wuhan-Hu1 variant were measured before (average, 202 days post symptom onset; Table S1), and 5-29 days (Table S1) after the first and second immunizations in the PIDs, and 6-28 days after the second immunization in the NDs. Three PIDs experienced asymptomatic SARS-CoV-2 infection (donors D, L, and M; Table S1) two of whom, L and M, did not have detectable anti-RBD IgG antibodies prior to immunization, while the third, D, had low but detectable serum anti-RBD IgG antibody titers (Fig. 2A). In the 13 PIDs with RBD-specific IgG antibodies prior to vaccination, a single dose of either vaccine boosted these titers by ∼500-fold (Fig. 2A). Across all PIDs there was a 200-fold increase in median RBD-specific IgA titers post-vaccination (Fig. 2B). Overall, in PIDs, a single vaccine dose elicited 4.5-fold higher IgG, and 7.7-fold higher IgA titers than two vaccinations in NDs. RBD-specific IgM titers were generally lower and were not significantly boosted in response to vaccination in both PIDs and NDs (Fig. 2C). In PIDs, a concomitant increase in RBD- (Fig. 2D) and S-specific IgG^+^ (Fig. 2E) memory B cell frequencies took place after vaccination. The two PIDs that lacked RBD-specific IgG titers prior to immunization (donors L and M) also lacked RBD-specific IgG+ memory B cells (Fig. 2D) and had lower frequencies of S-specific IgG^+^ memory B cells after vaccination. Consistent with the serology data, an increase in the frequency of IgA^+^ (Fig. 2F) but not IgM^+^ spike-specific memory B cells was observed (Fig. S3). Vaccination also induced S-specific CD4^+^ T-cell responses (Fig. 2G).

**Figure 2.**
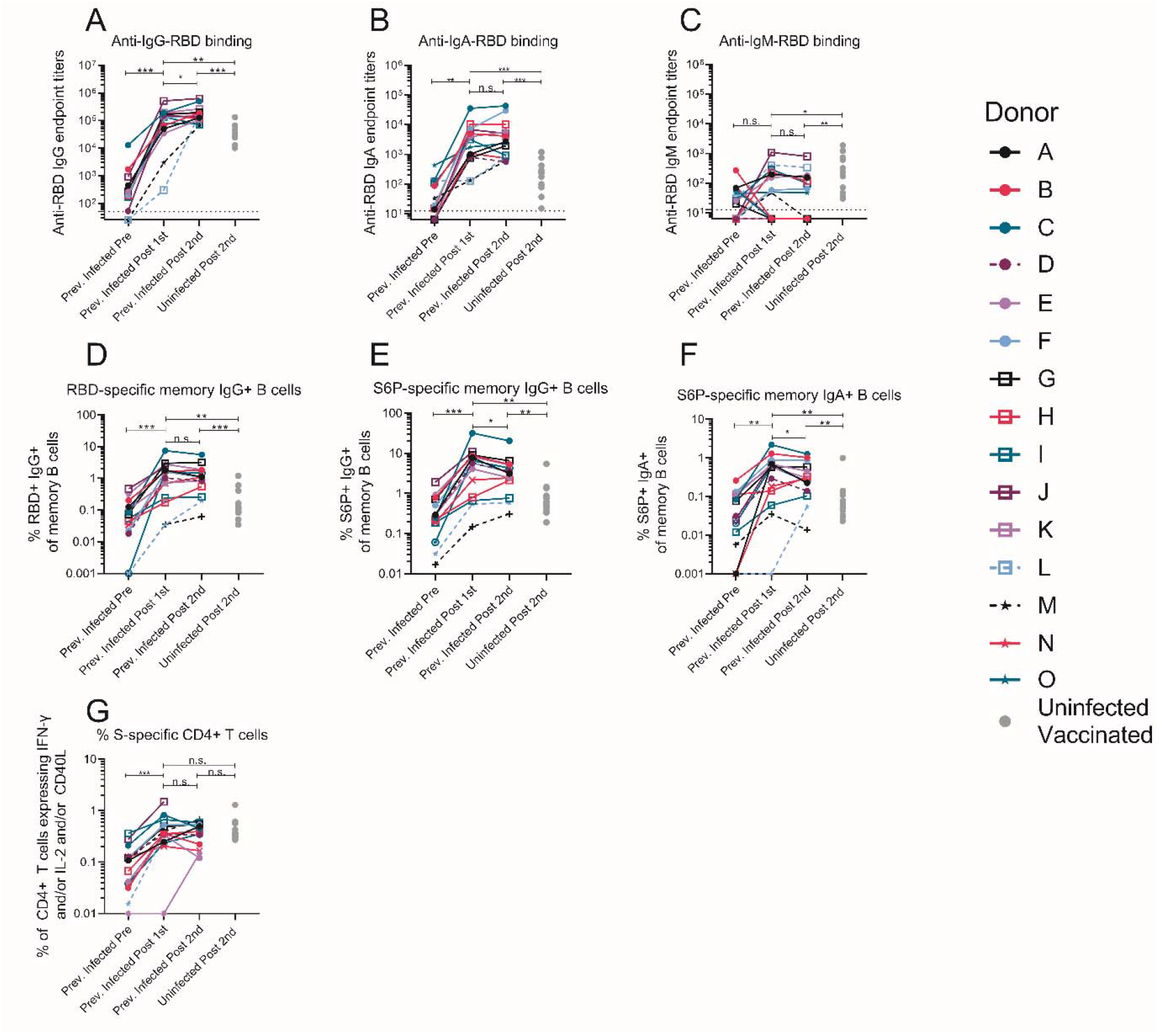
A single dose of a spike-derived mRNA vaccine elicits a strong recall response. IgG (**A**), IgA (**B**) and IgM (**C**) end-point antibody titers specific to the receptor binding domain of the Wuhan-Hu-1 variant were measured in serum collected from donors previously infected with SARS-CoV-2 before and after one or two immunizations with the Pfizer/BioNTech or Moderna mRNA vaccines by ELISA, as indicated. Endpoint titers measured in sera from uninfected donors following two vaccine doses are shown for comparison (gray dots). (**D**) Frequency of Wuhan-Hu-1 RBD-specific IgG+ memory B cells (live, IgD^-^, CD19^+^, CD20^+^, CD3^-^, CD14, CD56^-^, singlet, lymphocytes) in PBMC from previously infected donors was measured before and after one or two immunizations. The frequency of S6P-specific IgG^+^ (**E**) and IgA^+^ (**F**) memory B cells in PBMC previously infected donors were measured before and after one or two immunizations.The frequency of memory B cells from uninfected donors following two vaccine doses are shown for comparison in **D**-**F** (gray dots). (**G**) The frequency of S-specific CD4+ T cells expressing IFN-γ and/or IL-2 and/or CD40L in PBMC from previously infected donors was measured before and after one or two immunizations. The frequency of S-specific CD4+ T cells in PBMC from uninfected donors following two vaccine doses are shown for comparison (gray dots). Experiments were performed once. Significant differences in infected donors before or after vaccination (**A**-**D**) or between pseudoviruses (**E**) were determined using a Wilcoxon signed rank test (*p<0.05, **p<0.01 and ***p<0.001). Significant differences between previously infected and uninfected donors (**A**-**D**) were determined using a Wilcoxon rank sum test (*p<0.05, **p<0.01 and **p<0.001).

Sera from 12 of 15 PIDs sampled before vaccination neutralized the Wuhan-Hu-1 SARS-CoV-2 variant (Fig. 3A and S4). The non-neutralizing sera were from the three asymptomatic PIDs who had low or undetectable anti-RBD IgG titers (Fig. 3A dashed lines and Fig. S4), while pre-vaccine sera from the NDs were also non-neutralizing (Fig. S5). Consistent with the observed increase in binding antibodies following a single immunization in PIDs with pre-existing RBD-specific IgG titers, the median half-maximal neutralizing titers (ID_50_) were boosted approximately 1000-fold after the first dose, while the second dose had no effect (Fig. 3A). In the two PIDs lacking RBD-specific IgG titers prior to vaccination, the first vaccine dose elicited lower neutralizing titers (ID_50_ = ∼100 in donor L and ∼1,100 in donor M; Fig. 3A). In the NDs, two doses of the vaccine elicited ID_50_ titers that were ∼10- and 5-fold lower than those elicited by one or two doses in the PIDs, respectively (Fig. 3A and S6). Collectively, these data indicate that in PIDs who generate adequate immunological memory to the RBD, a single vaccine dose elicits an anamnestic response resulting in RBD-binding and neutralizing antibody responses that are superior to a two-dose regimen in uninfected donors.

**Figure 3.**
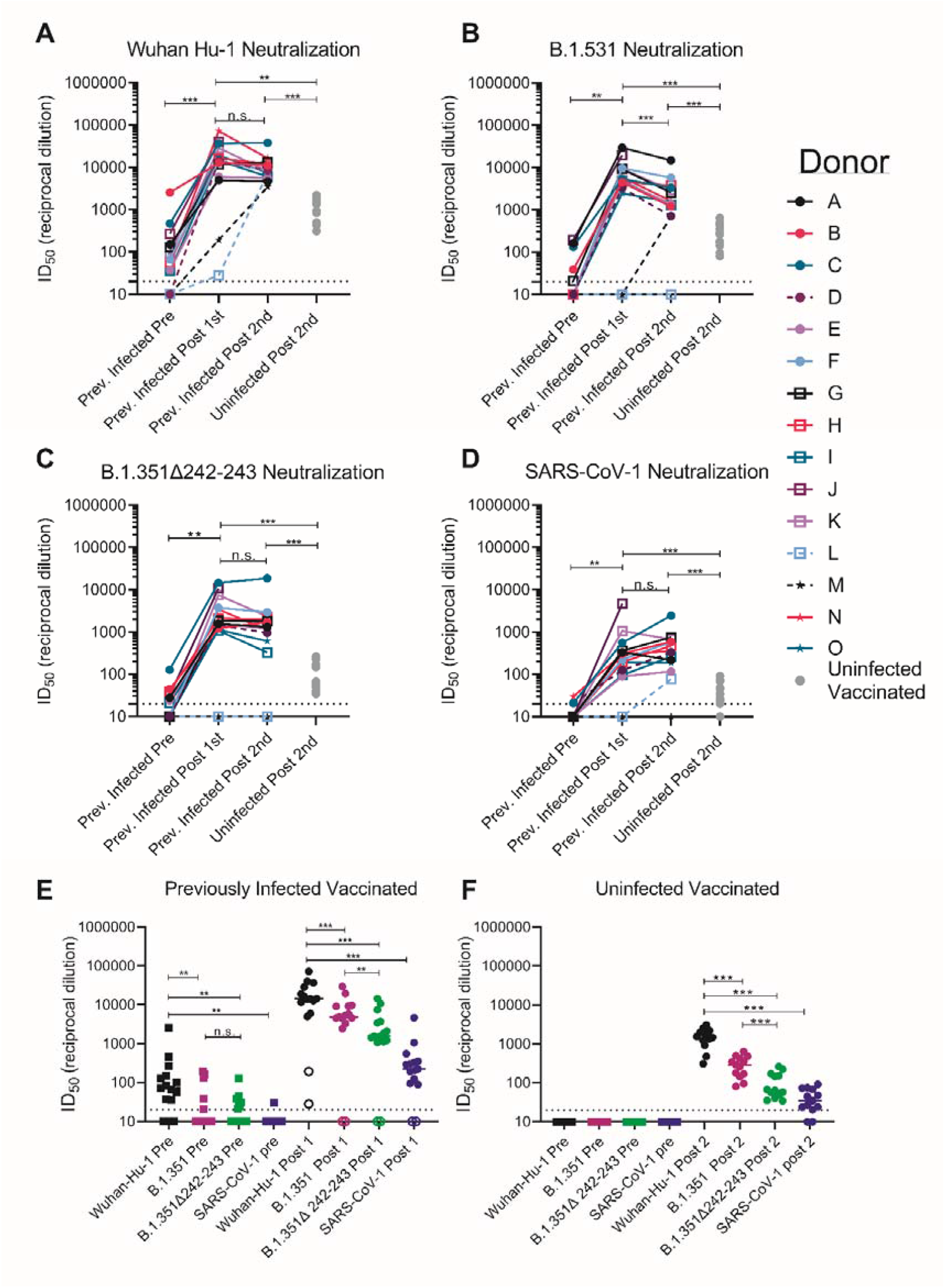
Pre-existing SARS-CoV-2 neutralizing antibody responses are boosted by a single dose of a spike-derived mRNA vaccine. The serum dilution resulting in 50% neutralization (ID_50_) of (**A**) Wuhan-Hu-1, (**B**) B.1.351, (**C**) B.1.351Δ242-243, and (**D**) SARS-CoV-1 pseudoviruses was measured in recovered COVID-19 donors prior to and following a one or two immunizations with the Pfizer/BioNTech or Moderna vaccines, and in uninfected donors following two vaccine doses as indicated. Data points between previously infected donors who were symptomatic and asymptomatic are connected by solid and dashed lines, respectively in **A**-**D. (E)** Serum dilution resulting in 50% neutralization (ID_50_) from recovered donors prior to (squares) and following a single immunization (circles) with the Pfizer/BioNTech or Moderna vaccines against Wuhan-Hu-1, B.1.351, B.1.351Δ242-243 and SARS-CoV-1 pseudoviruses as indicated. Previoulsy infected donors who were asymptomatic, negative for anti-IgG RBD antibodies, and RBD-specific IgG+ memory B cells prior to vaccination are shown as open circles. (**F**) Neutralizing potency (ID_50_) of serum from uninfected donors following two immunizations with the Pfizer/BioNTech or Moderna vaccines against the indicated pseudoviruses. Each data point represents a different donor and the horizonal bars represent the medians in **E** and **F**. The dashed lines demarcate the lowest serum dilutions tested. Experiments were performed once. Significant differences in infected donors before or after vaccination, or from the same timepoint against different variants (*p<0.05, **p<0.01 and ***p<0.001) were determined using a Wilcoxon signed rank test. Significant differences between previously infected and uninfected donors (*p<0.05, **p<0.01 and ***p<0.001) were determiend using a Wilcoxon rank sum test.

We next evaluated the ability of sera collected before and after immunization in NDs and PIDs to neutralize the more resistant B.1.351 and B.1.351-Δ242-243 pseudoviruses. These variants are 0.5% and 0.7% divergent from the Wuhan-Hu-1 variant. We also included SARS-CoV-1 pseudoviruses in this analysis, as a representative variant that is even more dissimilar to the vaccine. SARS-CoV-1 and SARS-CoV-2 are 24%, 26% and 50% divergent in the overall S protein, RBD and receptor binding motif, respectively (*80*). As a consequence, several mAbs that potently neutralize SARS CoV-2 fail to bind SARS-CoV-1 (*16, 22, 24*).

Prior to vaccination, 5 of 15 sera from PIDs neutralized B.1.351 and only three had ID_50_ titers above 100 (Figs. 3B and E and S4), 7 of 15 neutralized B.1.351Δ242-243, and only one had titers above 100 (Fig. 3C and E and S4). Only two pre-vaccine PID sera achieved 80% neutralization of B.1.351, and only one achieved 80% neutralization of B.1.351-Δ242-243 (Fig. S7A). The median ID_50_ of the pre-vaccine sera against Wuhan Hu-1 was significantly higher than against B.1.351 or B.1.351-Δ242-243 (Fig. 3E). Consistent with the high level of sequence disparity, sera from only one PID showed very weak neutralizing activity towards SARS-CoV-1 prior to vaccination (Fig. 3D and E and S7).

A single immunization boosted the nAb titers against all three SARS-CoV-2 variants and SARS-CoV-1 in 13 of 15 PIDs (Fig. 3 A-D); however, the median ID_50_ titers were ∼3-fold lower against B.1.351, ∼10-fold lower against B.1.351-Δ242-243, and 100-fold lower against SARS-CoV-1 than Wuhan-Hu-1 (Fig. 3E). A single immunization did not elicit nAbs against the B.1.351 variants or SARS CoV-1 in the two asymptomatic donors who lacked RBD-specific IgG memory (donor L and M; Fig. 3 A-D, and Fig. 3E open circles). The median ID_80_ values were also lower for the B.1.351 and B.1.351-Δ242-243 as compared to Wuhan-Hu-1 (Fig. S7A).

The neutralizing titers elicited by a single immunization in PIDs were significantly higher than those elicited by two immunizations in NDs against all pseudoviruses tested; 10-fold higher against Wuhan-Hu-1 (Fig. 3A), 20-fold higher against B.1.351 (Fig. 3B), 30-fold higher against B.1.351-Δ242-243 (Fig. 3C), and 7-fold higher against SARS-CoV-1 (Fig. 3D). Only 8 of 13 vaccinated NDs were able to achieve 80% neutralization of B.1.351-Δ242-243, and none could achieve 80% neutralization of SARS-CoV-1 (Fig. S7B).

The B.1.351 and B.1.351-Δ242-243 variants contain three RBD mutations that affect the neutralization potency of anti-RBD mAbs (Fig. 1). Moreover, pre-existing anti-RBD IgG memory appears to be important for a robust recall response to vaccination. To determine the relative contribution of anti-RBD antibodies to serum neutralization, we depleted RBD-specific antibodies from the sera of ten PIDs following one vaccination and from nine NDs after two vaccinations. This approach efficiently removed RBD-specific (Fig. 4A and C) but not anti-S2P specific antibodies from sera, as measured by ELISA (Fig. B and D). This depletion abrogated serum neutralization of Wuhan-Hu-1 virus (Fig. 4C and F), and B.1.351, B.1.351-Δ242-243, or SARS-CoV-1 (not shown), suggesting that the majority of neutralizing antibodies elicited or boosted by vaccination target this subdomain.

**Figure 4.**
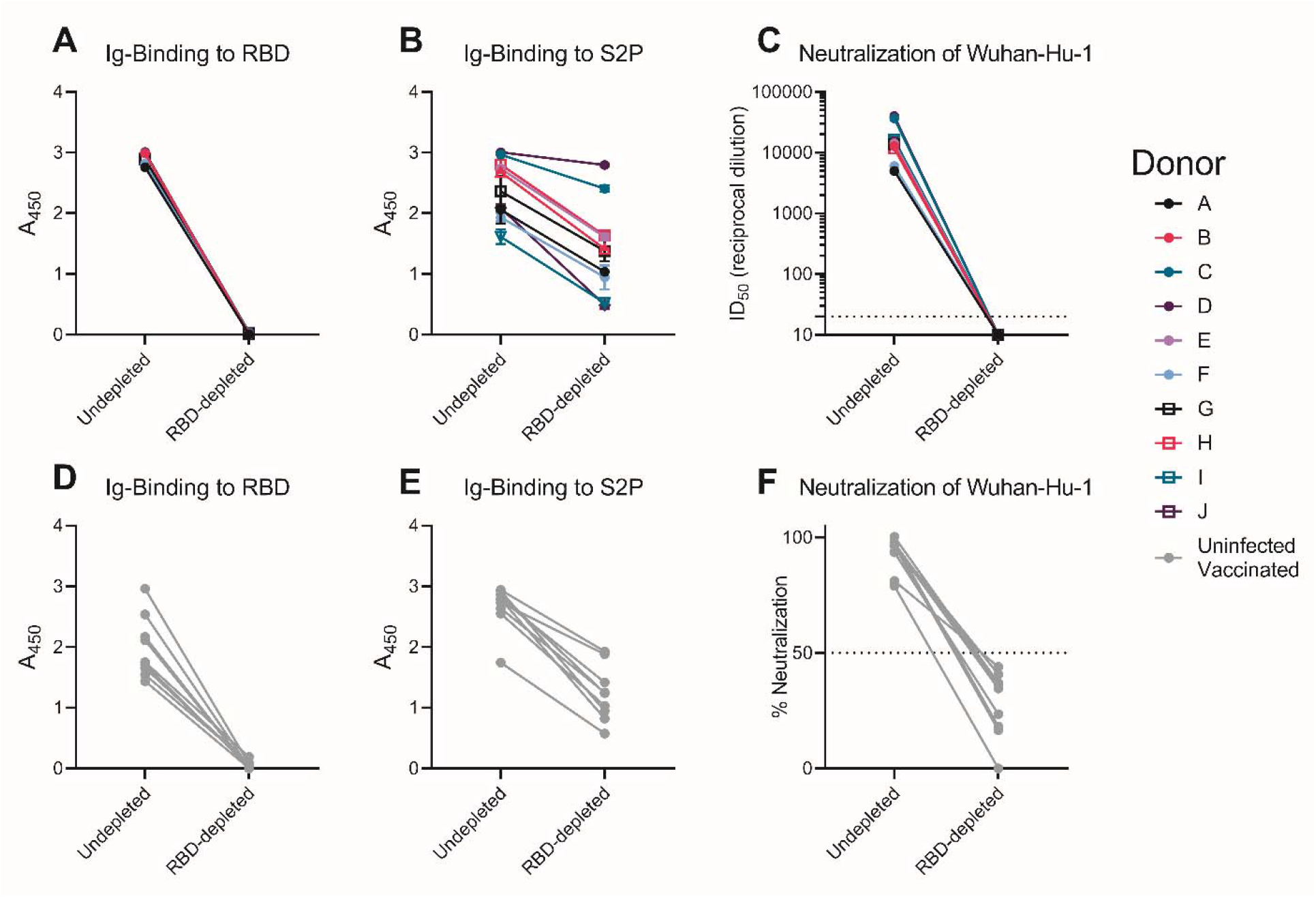
Vaccine-elicited neutralizing antibodies target the RBD. RBD-binding antibodies were adsorbed from sera from previously infected donors after receiving a single vaccine dose, or from uninfected donors after receiving two vaccine doses using Wuhan-Hu-1 RBD immobilized to magnetic beads. Antibody binding in undepleted or RBD-depleted serum from previously infected donors was measured to RBD at a 1:500 dilution (**A**), and S2P at a 1:4500 dilution (**B**) by ELISA as indicated. (**C**) The serum dilution resulting in 50% neutralization (ID_50_) of the Wuhan-Hu-1 pseudovirus was measured in undepleted or RBD-depleted of serum from the previously SARS-CoV-2 infected donors in **A** and **B**. Antibody binding in undepleted and RBD-depleted and sera from uninfected vaccinated donors was measured to RBD at a 1:500 dilution (**D**), and S2P at a 1:500 dilution (**E**) by ELISA. (**F**) The percent neutralization of a 1:120 dilution of undepleted or RBD-depleted of serum from the donors in D and E was measured against the Wuhan-Hu-1 pseudovirus. Experiments were performed once.

The above results indicate that in NDs, two doses of either the Pfizer/BioNTech or Moderna vaccines elicited nAb titers against the vaccine matched Wuhan-Hu-1, lower titers against B.1.351 and even lower titers against B.1.351-Δ242-243. Reduced sensitivity to vaccine-elicited nAbs has been reported for other B.1.351 variants (*66, 81, 82*).

Similarly, sera from PIDs who experienced symptomatic SARS-CoV-2 infection and who had detectable anti-RBD IgG titers prior to vaccination displayed generally weak nAb titers against Wuhan-Hu-1 at one to nine months post-infection and lower, or non-existent titers against the B.1.351 variants, in agreement with another report (*69*). However, provided RBD-specific IgG^+^ memory B-cell and antibody responses were generated during infection, a single immunization with either mRNA vaccine elicited a robust recall response that boosted the autologous neutralizing titers by approximately 1000-fold and importantly, these antibody responses cross-neutralized the B.1.351 variants, but at lower titers. In the majority of previously infected vaccinees, the anti-B.1.351-Δ242-243 neutralizing titers were comparable to those against the vaccine-matched Wuhan-Hu-1 in uninfected vaccinees. This is notable as these titers were associated with 95% protection from COVID-19 in phase 3 trials (*44, 46, 48, 49*). Moreover, vaccine-elicited antibody responses also neutralized SARS-CoV-1, but with much lower potencies. Collectively, our data suggest that the two mRNA vaccines that are based on the Wuhan-Hu-1 variant can elicit and/or boost neutralizing antibody responses, but that their potency is reduced against divergent variants.

Here, we show that the cross-neutralizing antibody responses generated after immunization in previously infected subjects are due to anti-RBD antibodies. Combined with the observation that the vaccines elicited neutralizing antibody responses that are less potent against the B.1.351 variant with the 242-243 deletion in the NTD suggests that NTD mutations can modulate the sensitivity of emerging variants to anti-RBD neutralizing antibodies. In contrast, the NTD region itself, which appears to tolerate antigenic variation in SARS-CoV-2 and other coronaviruses (*50, 52, 55, 83*), does not appear to be the target of cross-neutralizing antibodies elicited by infection or vaccination. We note that there are other less-frequent mutations associated with this lineage, such as L18F, Δ244, L244H, and R246I that were not examined here which may further increase resistance to vaccine-elicited antibodies.

Although the correlates of protection for SARS CoV-2 vaccines have not been established, studies in non-human primates indicate that even low titers of neutralizing antibodies are sufficient to prevent experimental SARS-CoV-2 infection, particularly if CD8+ T-cell responses are mounted (*18*). Our study suggests that most previously infected subjects will benefit from a single immunization with either the Pfizer/BioNTech or Moderna vaccines as it will lead to significant increases in serum neutralizing antibody responses against vaccine-matched and emerging variants. The observation that a second dose administered three to four weeks following the first in previously infected donors who have clear evidence of RBD-directed immunological memory prior to vaccination did not further boost neutralizing titers, suggests that the second dose of an mRNA vaccine could be delayed in some persons who have previously been infected with SARS-CoV-2. Longitudinal monitoring of the neutralizing antibody titers before and following the first dose should be used to determine the necessity, or optimal timing of the second dose in the context of previous infection.

## Materials and Methods

### Human Subjects

Peripheral blood mononuclear cells (PBMCs) and serum were collected from participants enrolled in the longitudinal study, “Seattle COVID-19 Cohort Study to Evaluate Immune Responses in Persons at Risk and with SARS-CoV-2 Infection”. Eligibility criteria included adults in the greater Seattle area at risk for SARS-CoV2 infection or those diagnosed with COVID-19 by a SARS CoV-2 PCR assay University of Washington multiplex PCR reaction using CDC approved primers/probes for the virus nucleocapsid (N) gene, N1 and N2) or blood antibody test (SARS CoV-2 IgG Architect, Abbot). Fifteen participants who recovered from SARS-CoV-2 and 13 that were SARS-CoV2 negative were selected from among those that were the first to receive two doses of an Emergency Use Authorization COVID-19 vaccine. Samples were selected based on availability. Sample sizes, randomization, or blinding were not pre-determined, nor were there specific inclusion/exclusion criteria. Serum or plasma from pre-pandemic controls were blindly selected at random from the study, “Establishing Immunologic Assays for Determining HIV-1 Prevention and Control”, with no considerations made for age, or sex. Both studies were recruited at the Seattle Vaccine Trials Unit (Seattle, Washington, USA). Informed consent was obtained from all participants and the Fred Hutchinson Cancer Research Center Institutional Review Board approved the studies and procedures (IR10440 and IR5567). Study data were collected and managed using REDCap electronic data capture tools hosted at Fred Hutchinson Cancer Research Center (*84, 85*).

### Cell Lines

All cell lines were incubated at 37°C in the presence of 5% CO2. 293-6E (human female, RRID:CVCL_HF20) and 293T cells (human female, RRID:CVCL_0063) cells were maintained in Freestyle 293 media with gentle shaking. HEK-293T-hACE2 (human female, BEI Resources Cat# NR-52511) were maintained in DMEM containing 10% FBS, 2 mM L-glutamine, 100 U/ml penicillin, and 100 µg/ml streptomycin (cDMEM).

### Recombinant CoV proteins and mAbs

A stabilized version of the recombinant SARS-CoV-2 spike protein (S2P) and the SARS CoV-2 RBD were produced in 293E cells as previously described (*8, 22*). Recombinant CV1, CV30 and AMMO1 were expressed in 293 cells and purified using protein A resin as previously described (*22, 79*). CV3-1 (Genbank: variable heavy region-MW681558, variable light region - MW681586), and CV2-75 (Genbank: variable heavy region-MW681758, variable light region - MW68175) mAbs were isolated from donors previously infected with SARS-CoV-2 using recombinant S2P as bait between 3.5 and 6 weeks post symptom-development, using methods outlined in (*22*).

### Generation of plasmids expressing SARS-CoV-1 and SARS-CoV-2 spike variants

To generate a plasmid encoding the SARS-CoV-2 spike B.1.351 variant (pHDM-SARS-CoV-2-Spike-B.1.351) we designed primers that anneal 5’ of the D80 codon and just 3’ of the A701 codon on the pHDM-SARS-CoV-2 Spike Wuhan-Hu-1 plasmid (BEI Resources Cat# NR-52514) that were used to amplify cDNA corresponding to the N and C termini of the spike protein and the plasmid backbone using Platinum SuperFi II DNA Polymerase (Thermofisher Cat# 12368010) according to the manufacturer’s instructions. cDNA encoding the rest of the spike protein including the D80A, D215G, K417N, E484K, N501Y, D614G and A701V mutations was synthesized as two g blocks (Integrated DNA technologies). The first had 30nt of homology with the PCR amplified vector backbone at the 5’ end. The second included and 30nt of homology with the 3’ end of the first block at the 5’ end and 30nt of homology with the PCR amplified vector backbone at the 3’ end. The g blocks and PCR product were ligated together using InFusion HD cloning Plus (TakaraBio Cat#638920). pHDM-SARS-CoV-2-Spike-B.1.351Δ242-243 was generated by deleting the amino acids 242-243 in pHDM-SARS-CoV-2-Spike-B.1.351 using site directed mutagenesis.

To generate a plasmid encoding the SARS-CoV-2 spike, codon-optimized cDNA corresponding to the SARS-CoV S protein (Urbani, GenBank: AAP13441.1) flanked on the 5’ end by 30nt of homology upstream of and including the EcoRI site, and flanked on the 3’ end by 30nt of homology downstream of and including the HindIII site on the pHDM-SARS-CoV-2 Spike Wuhan-Hu-1 plasmid was synthesized by Twist Biosciences. The synthesized DNA was cloned into the pHDM-SARS-CoV-2 Spike Wuhan-Hu-1 plasmid that was cut with EcoRI and HindIII and gel purified to remove the SARS-CoV-2 Spike cDNA using InFusion HD cloning Plus. The sequences of the cDNA in pHDM-SARS-CoV-Spike-B.1.351 and pHDM-SARS-CoV Spike were verified by Sanger sequencing (Genewiz Inc.).

### Pseudovirus neutralization assay

HIV-1 derived viral particles were pseudotyped with full length wildtype S from Wuhan Hu1, B.1.351, B.1.351Δ242-243, or SARS CoV-1 using a previously described reporter system (*86*), Briefly, plasmids expressing the HIV-1 Gag and pol (pHDM540 Hgpm2, BEI Resources Cat# NR-52517), HIV-1Rev (pRC-CMV-rev1b, BEI Resources Cat# NR-52519), HIV-1 Tat (pHDM-tat1b, BEI resources NR-52518), the SARS-CoV-2 spike (pHDM-SARS-CoV-2 Spike Wuhan-Hu-1, pHDM-SARS-CoV-2 Spike B.1.351, or pHDM-SARS-CoV-1 Spike) and a luciferase/GFP reporter (pHAGE-CMV-Luc2-IRES542 ZsGreen-W, BEI Resources Cat# NR-52516) were co-transfected into 293T cells at a 1:1:1:1.6:4.6 ratio using 293 Free transfection reagent according to the manufacturer’s instructions. Pseudoviruses lacking a spike protein were also produced as a control for specific viral entry. Pseudoviron production was carried out at 32 °C for 72 hours after which the culture supernatant was harvested, clarified by centrifugation and frozen at −80°C.

293 cells stably expressing human HEK-293T-hACE2 were seeded at a density of 4×10^3^ cells/well in a 100 µL volume in 96 well flat bottom black-walled, clear bottom tissue culture plates (Greiner CELLSTAR Cat#655090). The next day, mAbs or sera were serially diluted in 70 µL of cDMEM in 96 well round bottom master plates in duplicate wells. 30 µL of serially diluted mAbs or sera from the master plate were replica plated into 96 well round bottom plates. An equal volume of viral supernatant diluted to achieve comparable luciferase values between SARS-CoV-2 variants (Fig S1) was added to 96 well round bottom plates containing identical serial dilutions from the same master plate, and incubated for 60 min at 37 °C. Meanwhile 50 μL of cDMEM containing 6 µg/mL polybrene was added to each well of 293T-ACE2 cells (2 µg/mL final concentration) and incubated for 30 min. The media was aspirated from 293T-ACE2 cells and 100 µL of the virus-antibody mixture was added. The plates were incubated at 37°C for 72 hours. The supernatant was aspirated and replaced with 100 μL of Steadyglo luciferase reagent (Promega Cat# E2510) and read on a Fluorskan Ascent Fluorimeter. Control wells containing virus but no antibody (cells + virus) and no virus or antibody (cells only) were included on each plate.

Percent neutralization for each well was calculated as the RLU of the average of the cells + virus wells, minus test wells (cells +mAb/sera + virus), and dividing this result difference by the average RLU between virus control (cells + virus) and average RLU between wells containing cells alone, multiplied by 100. The antibody concentration, or serum dilution that neutralized 50% or 80% of infectivity (IC_50_ and IC_80_ for mAbs, ID_50_ and ID_80_ for serum) was interpolated from the neutralization curves determined using the log(inhibitor) vs. response -- Variable slope (four parameters) fit using automatic outlier detection in Graphpad Prism Software. Serum that did not achieve 50% neutralization at the lowest dilution tested (1:20) were considered negative and assigned an ID_50_ value of 10.

### RBD ELISAs

Half-well area plates (Greiner) were coated with purified RBD protein at 16.25ng/well in PBS (Gibco) for 14-24h at room temperature. After 4 150ul washes with 1X PBS, 0.02% Tween-2 (Sigma) using the BioTek ELx405 plate washer, the IgA and IgG plates were blocked at 37°C for 1-2 hours with 1X PBS, 10% non-fat milk (Lab Scientific), 0.02% Tween-20 (Sigma); IgM plates were blocked with 1X PBS, 10% non-fat milk, 0.05% Tween-20.

Serum samples were heat inactivated by incubating at 56°C for 30 minutes, then centrifuged at 10,000 x g / 5 minutes, and stored at 4°C previous to use in the assay. For IgG ELISAs, serum was diluted into blocking buffer in 7-12 1:4 serial dilutions stating at 1:50. For IgM and IgA ELISAs, serum was diluted into 7 1:4 serial dilutions stating at 1:12.5 to account for their lower concentration. A qualified pre-pandemic sample (negative control) and a standardized mix of seropositive serums (positive control) was run in each plate and using to define passing criteria for each plate. All controls and test serums at multiple dilutions were plated in duplicate and incubated at 37°C for 1 hour, followed by 4 washes in the automated washer. 8 wells in each plate did not receive any serum and served as blocking controls.

Plates then were plated with secondary antibodies (all from Jackson ImmunoResearch) diluted in blocking buffer for 1h at 37C. IgG plates used donkey anti-human IgG HRP Cat # 709-035-098) diluted at 1:7500; IgM plates used goat anti-human IgM HRP (Cat# 109-035-043) diluted at 1:10,000; IgA plates used goat anti-human IgA HRP (Cat# 109-035-011) at 1:5000. After 4 washes, plates were developed with 25ul of SureBlock Reserve TMB Microwell Peroxide Substrate (Seracare) for 4 min, and the reaction stopped by the addition of 50μl 1N sulfuric acid (Fisher) to all wells. Plates were read at OD_450nm_ on SpectraMax i3X ELISA plate reader within 20 min of adding the stop solution.

OD_450nm_ measurements for each dilution of each sample were used to extrapolate endpoint titers when CVs were less than 20%. Using Excel, endpoint titers were determined by calculating the point in the curve at which the dilution of the sample surpassed that of 5 times the average OD_450nm_ of blocking controls + 1 standard deviation of blocking controls.

### RBD-specific antibody depletion

SARS-CoV-2 RBD was coupled to Pierce NHS-activated magnetic beads (Thermo Fisher) at a ratio of 50 µg of RBD to 1 mg of beads according to manufacturer’s instructions and stored in PBS. Plasma samples were diluted 1:50 in DMEM in a total volume of 500µl. Diluted plasma was incubated with of 65 ul of RBD-coupled beads. Samples were continually mixed at RT for 1 hour. Diluted sera was removed from immobilized magnetic beads using a magnetic stand and the sera were incubated with fresh beads two additional times. Depleted serum was filter sterilized using a 0.22 um spin filter. The presence of binding antibodies in depleted and undepleted sera was measured using recombinant RBD and S2P by ELISA using a polyclonal anti-IgG secondary (Southern Biotech Cat# 2010-05)

### Spike and RBD memory B cell flow cytometry assays

Fluorescent SARS-CoV-2-specific S6P (*87*) (provided by Roland Strong, Fred Hutchinson Cancer Research Center, Seattle, WA) and RBD probes were made by combining biotinylated protein with fluorescently labeled streptavidin. The S6P probes were made at a ratio of 1:1 molar ratio of trimer to SA. Two S6P probes, one labeled with AlexaFluor488 (Invitrogen), one labeled with AlexaFluor647 (Invitrogen), were used in this panel in order to increase specificity of the detection of SARS-CoV-2-specific B cells. The RBD probe was prepared at a 4:1 molar ratio of RBD monomers to SA, labeled with R-phycoerythrin (Invitrogen). Cryopreserved PBMCs from SARS-CoV-2-convalescent participants and a pre-pandemic SARSCoV-2-naïve donor were thawed at 37°C and stained for SARS-CoV-2-specific memory B cells with a flow cytometry panel shown in Supplementary Table 3. Cells were stained first with the viability stain (Invitrogen) in PBS for 15 min at 4°C. Cells were then washed with 10% FBS/PBS and stained with a cocktail of the three probes for 30 min at 4°C. The probe cocktail was washed off with 10% FBS/PBS and the samples were stained with the remaining antibody panel and incubated for 25 min at 4°C. The cells were washed two times and resuspended in 1% paraformaldehyde/1x PBS for collection on an LSR II flow cytometer (BD Biosciences). Data was analyzed in Flow Jo version 9.9.4.

### Intracellular Cytokine Staining (ICS) Assay

Flow cytometry was used to examine SARS-CoV-2-specific CD4+ T-cell responses using a validated ICS assay. The assay was similar to published reports(*88, 89*) and the details of the staining panel are included in Supplemental Table 4. Two peptide pools (15 amino acids overlapping by 11 amino acids, provided by BioSynthesis) covering the spike protein of SARS-CoV-2 were used for the six-hour stimulation. Peptide pools were used at a final concentration of 1 µg/ml for each peptide. As a negative control, cells were not stimulated, only the peptide diluent (DMSO) was included. As a positive control, cells were stimulated with a polyclonal stimulant, staphylococcal enterotoxin B (SEB). Cells expressing IFN-γ and/or IL-2 and/or CD154 was the primary endpoint for antigen-specific CD4^+^ T cells. The overall response to spike was defined as the sum of the background-subtracted responses to the spike 1 and spike 2 individual pools. The total number of CD4^+^ T cells must have exceeded 10,000 the assay data to be included in the analysis.

### Biolayer interferometry (BLI)

BLI experiments were performed on an Octet Red instrument at 30°C with shaking at 500-1000 rpm. mAbs were loaded at a concentration of 20 mg/mL in PBS onto Anti-Human IgG Fc capture (AHC) biosensors (Fortebio) for 300s, followed by a 60s baseline in KB buffer (1X PBS, 0.01% Tween 20, 0.01% BSA, and 0.005% NaN_3_, pH 7.4). Probes were then dipped in KB containing either; SARS-CoV-2 RBD (2.0µM), S-2P (0.5µM), S1 NTD (0.5µM, SinoBiological Cat # 40591-V49H) or S2 (0.5µM, SinoBiological Cat # 40590-V08B) for a 300s association phase, followed by a 300s dissociation phase in KB. The binding of mature VRC01 (*90*) was used as negative control to subtract background binding at each timepoint.

## Data Availability

All data is provided in the manuscript.

## ACKNOWLEDGMENTS

This work was supported by generous donations to Fred Hutch COVID-19 Research Fund, and to MJM from the Paul G. Allen Family Foundation, the Joel D. Meyers Endowed Chair, and NIAID UM1 AI068618-14S1, 2UM1 AI069481-15, and UM1A057266-S1. We thank the study participants for their dedication to this project, Trevor Bedford for assistance with the selection of spike mutations to include, Laura Richert Spuhler for assistance with figure preparation, Todd Haight and the Seattle Vaccine Unit specimen processing lab and staff for their service, and. L.S. and A.T.M. have filed a provisional patent application on the CV1, CV2-75 and CV30 SARS-CoV-2 specific monoclonal antibodies. L.S., A.T.M., and A.F. have filed a provisional patent application on the CV3-1 SARS-CoV-2 specific monoclonal antibody.

## AUTHOR CONTRIBUTIONS

Conceptualization, M.J.M., A.T.M., and L.S.; Investigation, Y-H.W., E.S., M.L., V.R., K.W.C., Z.M., M.N., L.J.H., A.J.M., M.J, J.F, G.M., H.G., and A.T.M; Writing - Original Draft, A.T.M., and L.S.; Writing - Review & Editing, A.T.M., L.S., M.J.M., and E.S.; Funding Acquisition, L.S. and M.J.M; Resources, M.J.M., J.C., A.F.; Supervision, A.T.M.

## List of Supplementary Materials

Figs. S1 to S8

Tables S1 to S4

## Notes

### Competing Interest Statement

The authors have declared no competing interest.

### Author Declarations

Peripheral blood mononuclear cells (PBMCs) and serum were collected from donors who recovered from SARS-CoV-2 infection and were the first 10 who then subsequently received a SARS-CoV-2 vaccine as part of the study: Seattle COVID-19 Cohort Study to Evaluate Immune Responses in Persons at Risk and Peripheral blood mononuclear cells (PBMCs) and serum were collected from participants enrolled in the longitudinal study: Seattle COVID-19 Cohort Study to Evaluate Immune Responses in Persons at Risk and with SARS-CoV-2 Infection. Eligibility criteria included adults in the greater Seattle area at risk for SARS-CoV2 infection or those diagnosed with COVID-19 by a SARS CoV-2 PCR assay University of Washington multiplex PCR reaction using CDC approved primers/probes for the virus nucleocapsid (N) gene, N1 and N2) or blood antibody test (SARS CoV-2 IgG Architect, Abbot). Fifteen participants who recovered from SARS-CoV-2 and 13 that were SARS-CoV2 negative were selected from among those that were the first to receive two doses of an Emergency Use Authorization COVID-19 vaccine. Samples were selected based on availability. Sample sizes, randomization, or blinding were not pre-determined, nor were there specific inclusion/exclusion criteria. Serum or plasma from pre-pandemic controls were blindly selected at random from the study: Establishing Immunologic Assays for Determining HIV-1 Prevention and Control, with no considerations made for age, or sex. Both studies were recruited at the Seattle Vaccine Trials Unit (Seattle, Washington, USA). Informed consent was obtained from all participants and the Fred Hutchinson Cancer Research Center Institutional Review Board approved the studies and procedures (IR10440 and IR5567). Study data were collected and managed using REDCap electronic data capture tools hosted at Fred Hutchinson Cancer Research Center.

### Summary of Updates

This version is updated to include: -antibody-binding, immune cell frequency and neutralization data from additional donors with previous SARS-CoV-2 infection after one or two immunizations with mRNA vaccines. -antibody-binding, immune cell frequency and neutralization data from uninfected donors after two immunizations with mRNA vaccines -experimental evidence that the cross-neutralizing antibodies target the SARS-CoV2 receptor binding domain

