## Supplementary material for "A single mRNA immunization boosts cross-variant neutralizing antibodies elicited by SARS-CoV-2 infection": Figures S1-S8, Tables S1-S4

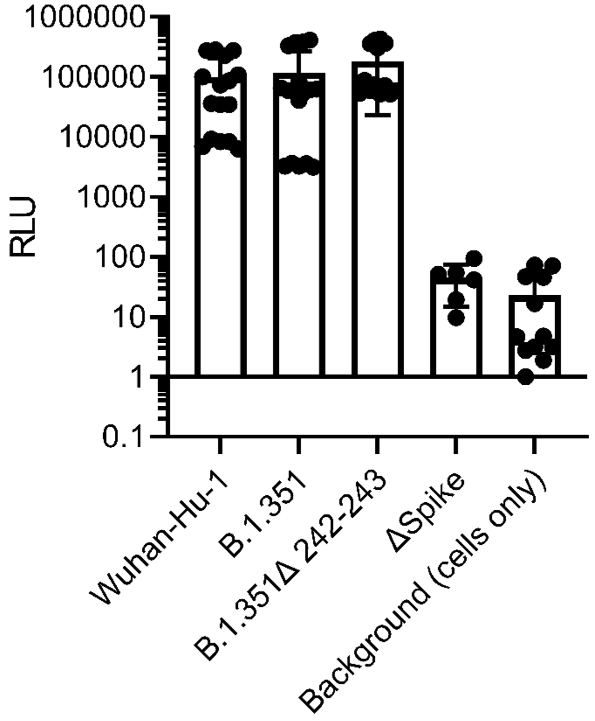


**Fig. S1.**

Viral dilutions yielding similar relative infectivity of Wuhan-Hu-1, B.1.351, and B.1.351Δ242-243 pseudoviruses in 293-ACE2 cells in the absence of antibody or sera were employed during neutralization experiments. Each dot represents the average entry of measured n=8 replicate wells from an individual assay plate corresponding to the data shown in Figure 3. The entry of pseudoviral particles produced without transfection with a spike-encoding plasmid are shown for comparison (Δspike). The background is the luciferase signal obtained from n=8 replicate wells to which neither virus was added. The bars represent the mean and the error bars represent the SD.

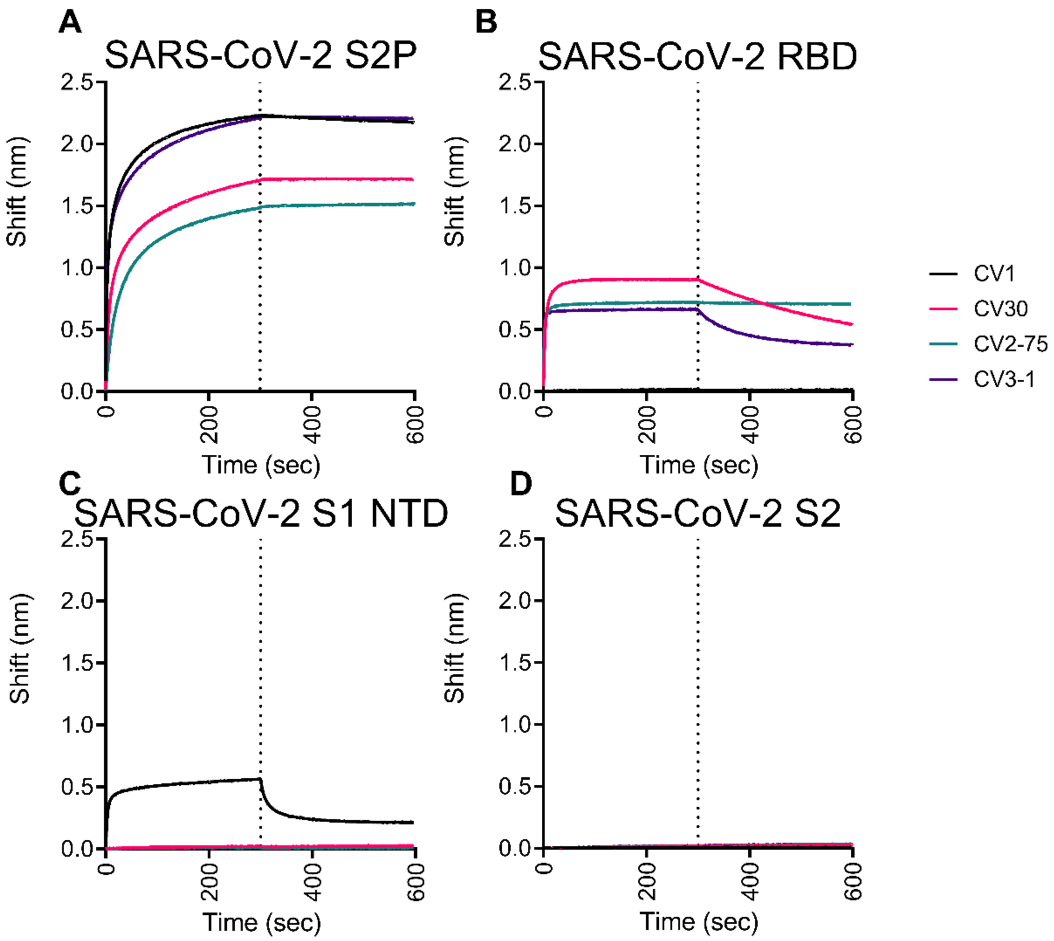


**Fig. S2.**

Binding of the indicated mAbs was measured to recombinant (A) SARS-CoV-2 stabilized spike (S2P), (B) SARS-CoV-2 receptor binding domain (RBD), (C) SARS-CoV-2 S1 N-terminal domain (NTD), and (D) SARS CoV-2 S2 (S2) by biolayer interferometry.

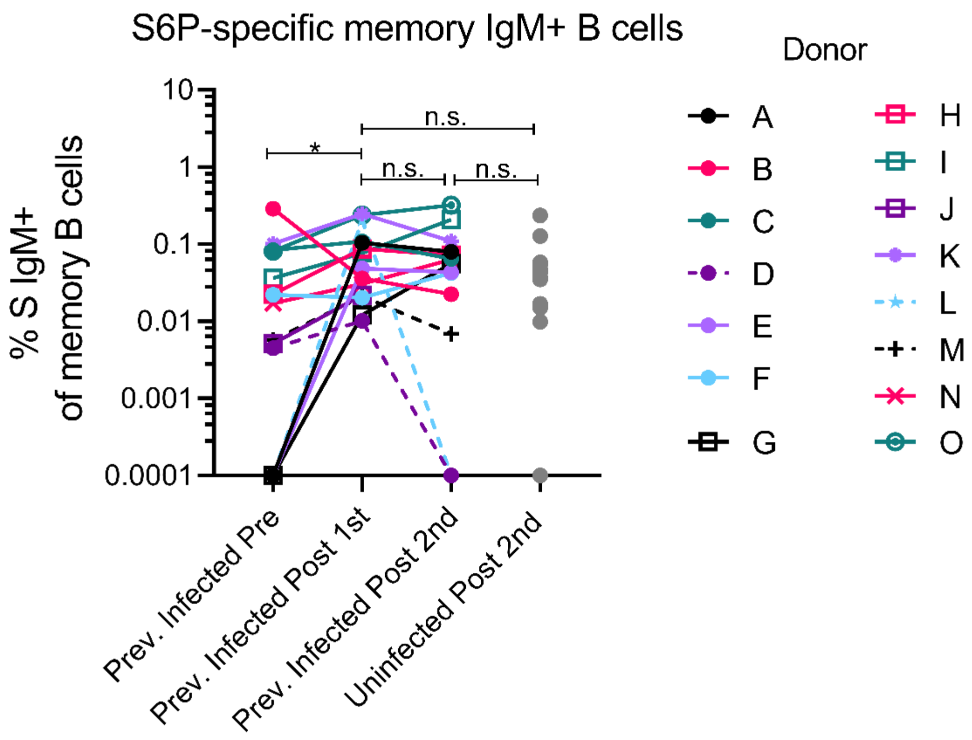


**Fig. S3.**

Frequency of stabilized SARS-CoV-2 S6P-specific IgM memory B cells (live, IgD-, CD19+, CD20+, CD3-, CD14-, CD56-, singlet, lymphocytes) from previously infected SARS-CoV-2-donors were measured prior to and following one or two immunizations with the Pfizer/BioNTech or Moderna vaccines. Data points between previously infected donors who were symptomatic and asymptomatic are connected by solid and dashed lines, respectively. IgM memory B cells were measured in uninfected donors who received two doses of either vaccine are included for comparison (gray dots). Experiment was performed once. Significant differences in infected donors before or after vaccination (* $p<0.5$) were determined using a Wilcoxon signed rank test. Significant differences between previously infected and uninfected donors were ruled out using a Wilcoxon rank sum test.

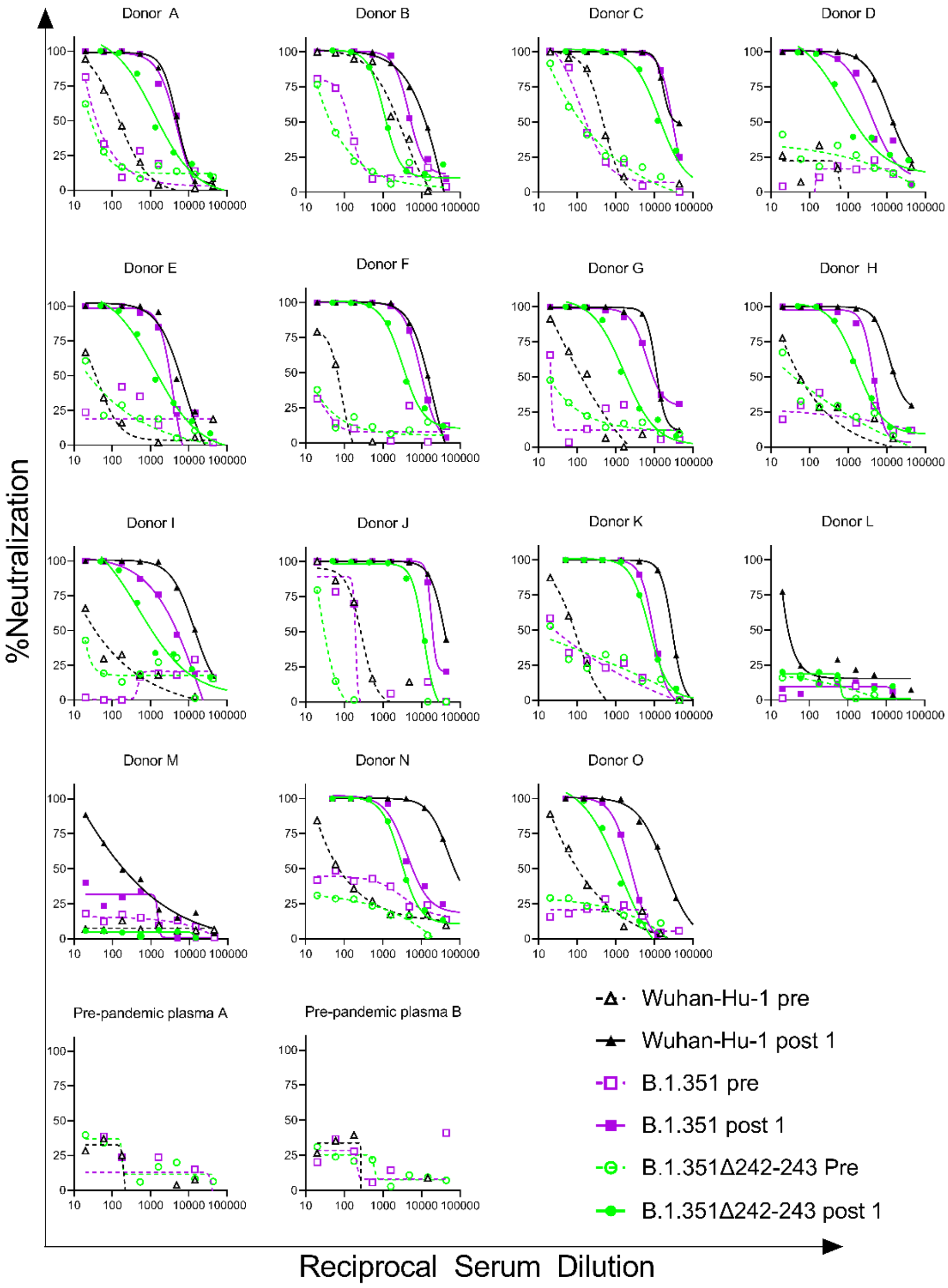


**Figure S4.**

The ability of sera from donors who were previously infected with SARS-CoV-2 were evaluated for their ability to neutralize the Wuhan-Hu-1, B.1.351, and B.1.351Δ242-243 pseudovirus infectivity before and after immunization as indicated. Plasma collected from two donors prior to the pandemic were included as controls. Data points represent the mean of two technical replicates.

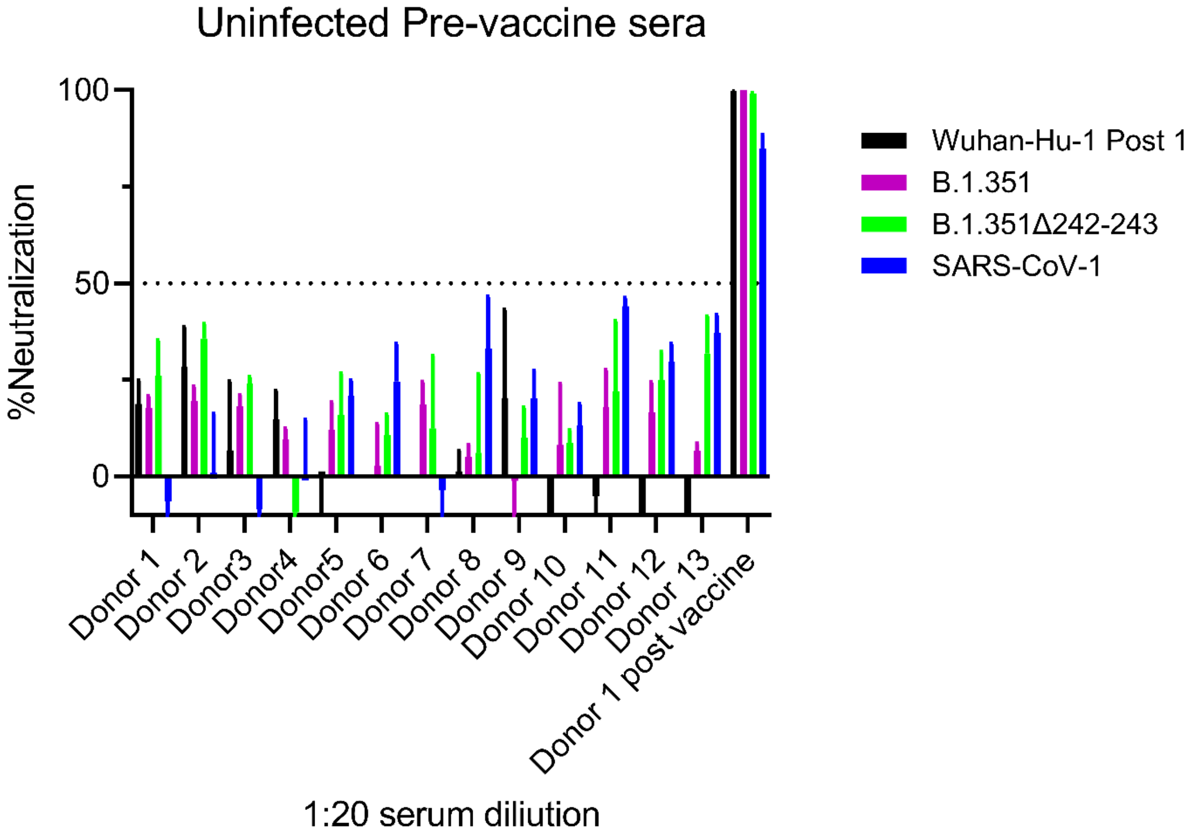


**Figure S5**.
Serum from SARS-CoV-2 uninfected donors collected prior to vaccination with the Pfizer/BioNTech or Moderna mRNA vaccines were diluted 1:20 and evaluated for their ability to neutralize the indicated SARS-CoV-2 variants and SARS-CoV-1 as indicated. Sera from donor 1 collected following two immunizations with the Pfizer/BioNTech vaccine is included as a control. Dashed line indicates the cutoff for neutralization. Bars represent the mean and error bars represent the SD of three technical replicates.

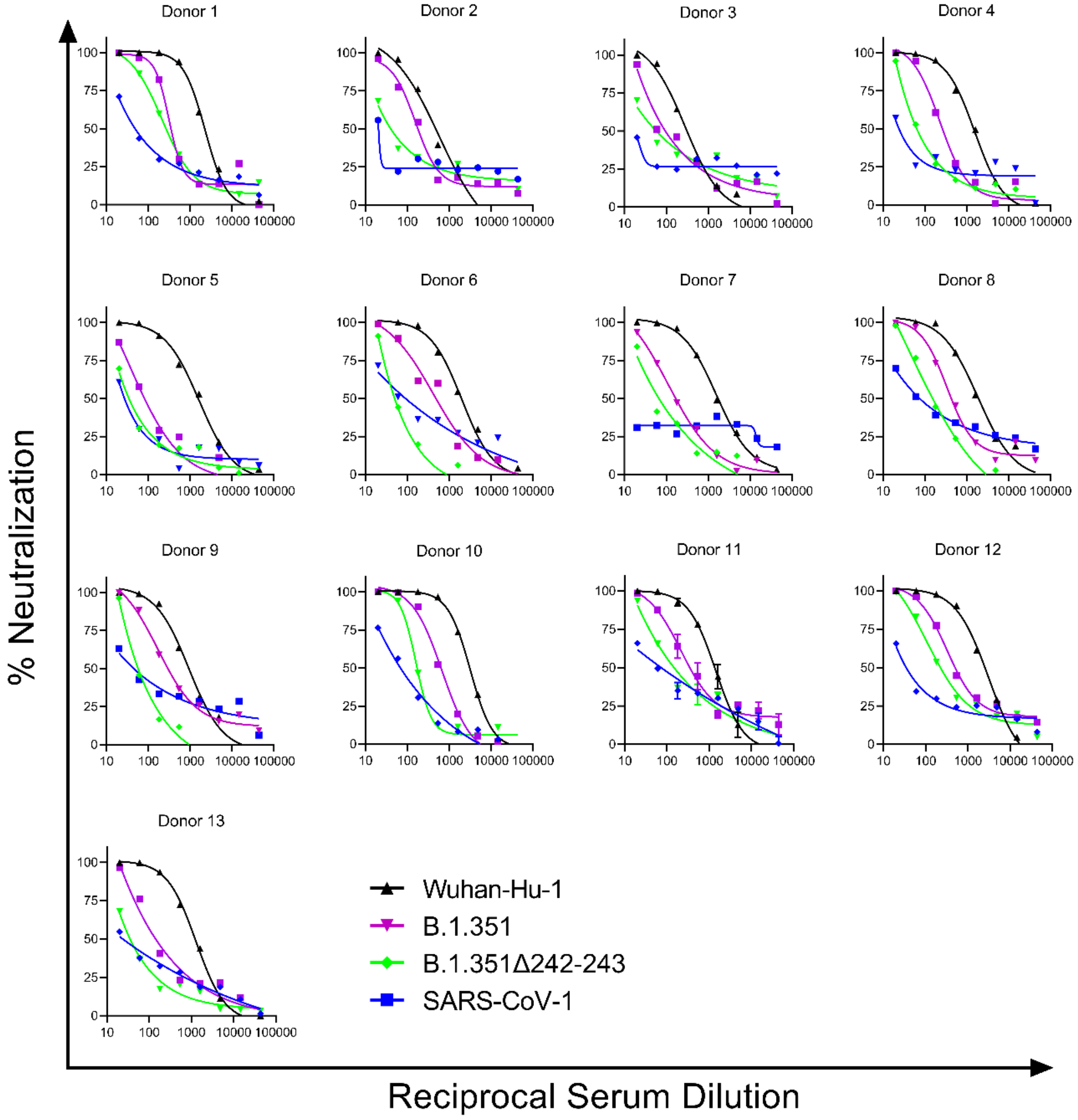


**Figure S6**.
The ability of serially diluted serum from previously uninfected donors following two immunizations with the Pfizer/BioNTech or Moderna vaccines was evaluated for their ability to neutralize the indicated pseudoviruses. Data points represent the mean of two technical replicates.

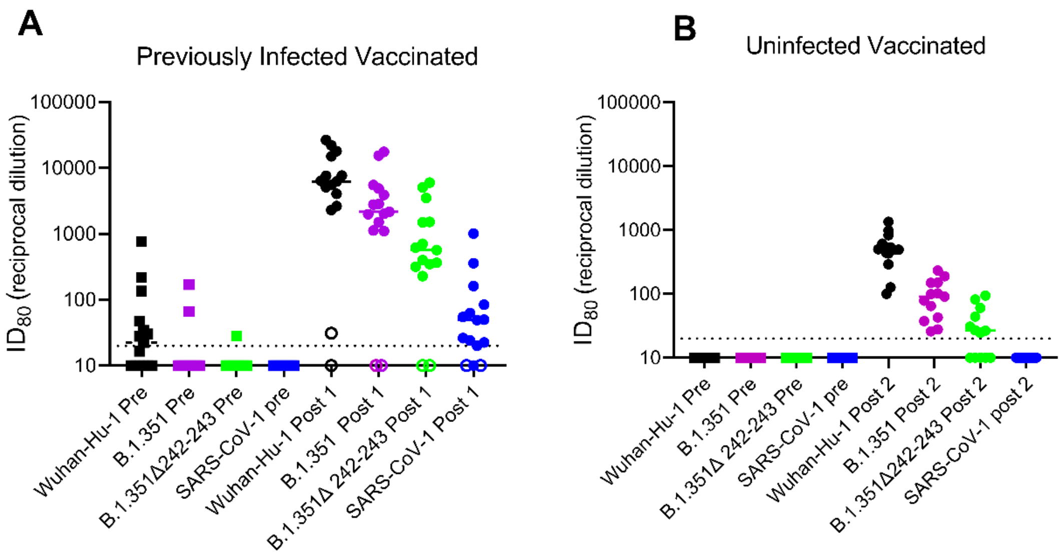


**Figure S7.**

**(A)** Serum dilution resulting in 80% neutralization ($ID_{80}$) from recovered donors prior to (squares) and following a single immunization (circles) with the Pfizer/BioNTech or Moderna vaccines against Wuhan-Hu-1, B.1351, B.1351Δ242-243 SARS CoV-2 pseudoviruses and SARS-CoV-1 pseudoviruses as indicated. Previoulsy infected donors who were asymptomatic, negative for anti-IgG RBD antibodies, and RBD-specific IgG+ memory B cells prior to vaccination are shown as open circles. (**B**) Neutralizing potency ($ID_{50}$) of serum from uninfected donors following two immunizations with the Pfizer/BioNTech or Moderna vaccines against the indicated pseudoviruses. Each data point represents a different donor and the horizonal bars represent the medians**.** The dashed lines demarcate the lowest serum dilutions tested.

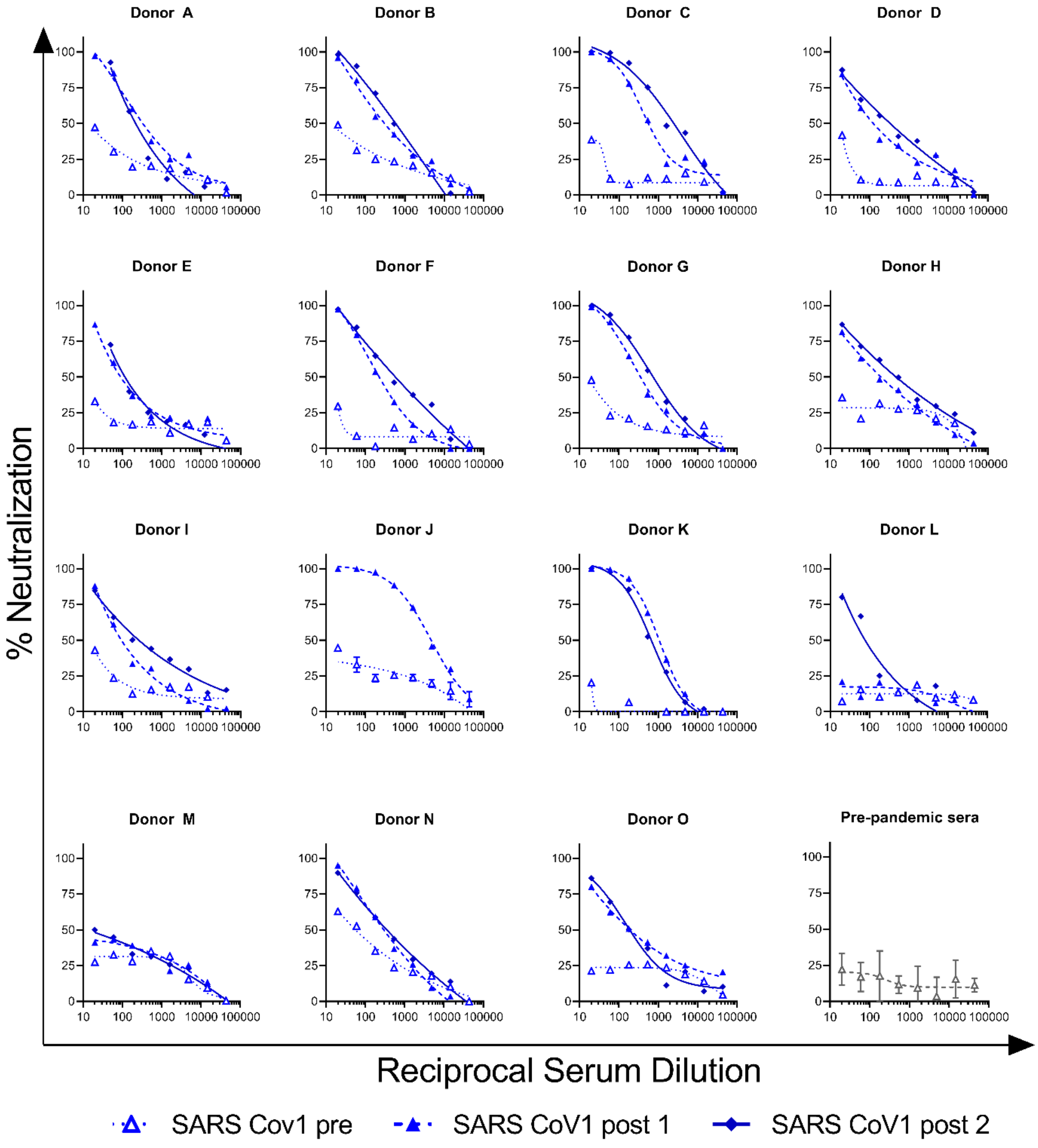


**Figure S8.**

The ability of serum from donors who recovered from SARS-CoV-2 were evaluated for their ability to neutralize SARS-CoV-1 infectivity before and after immunization as indicated. Data points represent the mean of two technical replicates. Serum collected from a donor prior to the pandemic was included as a control (bottom right panel).

**Table S1.**

**Seattle COVID-19 Study Population - previously infected.**

| Participant ID | Age (years) | Sex at Birth | Race or ethnicity | Disease severity (WHO scale) | Days from symptom onset to pre-vaccine visit | Days from pre-vaccine visit to vaccination | Days from first vaccine to post- 1 vaccination visit | Days from first vaccine to Second vaccine | Days from second vaccine to post- 2 vaccination visit | SARS-CoV-2 Vaccine |
|---|---|---|---|---|---|---|---|---|---|---|
| A | 50s | Male | White | 3 | 271 | 2 | 18 | 18 | 29 | Pfizer |
| B | 40s | Male | Hispanic / Latino | 2 | 31 | 243 | 16 | 21 | 16 | Pfizer |
| C | 50s | Male | White | 3 | 128 | 136 | 17 | 17 | 19 | Pfizer |
| D | 60s | Female | White | n/a* | n/a* | 71 | 14 | 21 | 7 | Pfizer |
| E | 50s | Male | White | 2 | 247 | 66 | 14 | 21 | 26 | Pfizer |
| F | 30s | Female | White | 2 | 202 | 76 | 18 | 18 | 18 | Pfizer |
| G | 50s | Female | White | 2 | 193 | 78 | 20 | 20 | 9 | Pfizer |
| H | 40s | Female | White | 3 | 237 | 12 | 13 | 28 | 6 | Moderna |
| I | 40s | Male | White | 2 | 64 | 24 | 16 | 27 | 5 | Moderna |
| J | 50s | Male | Asian | 3 | 274 | 15 | 13 | 31 | 10 | Moderna |
| K | 40s | Male | White | 2 | 270 | 6 | 16 | 27 | 10 | Moderna |
| L | 40s | Male | White | n/a* | n/a* | 14 | 14 | 22 | 13 | Pfizer |
| M | 40s | Male | White | n/a* | n/a* | 50 | 20 | 22 | 16 | Pfizer |
| N | 50s | Female | White | 2 | 115 | 27 | 15 | 20 | 10 | Pfizer |
| O | 40s | Female | White | 2 | 269 | 6 | 17 | 21 | 16 | Pfizer |
| **N=15 median (IQR) or %** | 49 (45-56) | 60% Male | 87% White | 27% WHO scale 3 | 220 (125-269) | 27 (13-74) | 16 (14-18) | 21 (20-24) | 13 (10-17) | 73% Pfizer |

*participants D, L and M were asymptomatic.
n/a: not available

**Table S2.**

Seattle COVID-19 Study Population-uninfected

| Participant ID | Age (years) | Sex at Birth | Race or ethnicity | Days from pre-vaccine visit to vaccination | Days from second vaccine to post-2 vaccination visit | SARS-CoV-2 Vaccine |
|---|---|---|---|---|---|---|
| 1 | 30s | Female | White | 244 | 19 | Pfizer |
| 2 | 50s | Male | Asian;White | 241 | 22 | Pfizer |
| 3 | 60s | Male | White | 251 | 8 | Pfizer |
| 4 | 40s | Male | White | 236 | 13 | Pfizer |
| 5 | 30s | Male | White | 224 | 19 | Pfizer |
| 6 | 20s | Male | Asian;White | 237 | 16 | Pfizer |
| 7 | 40s | Male | White | 240 | 18 | Pfizer |
| 8 | 50s | Male | White | 228 | 19 | Pfizer |
| 9 | 30s | Male | White | 150 | 28 | Pfizer |
| 10 | 50s | Male | White | 247 | 6 | Moderna |
| 11 | 40s | Male | White | 233 | 22 | Pfizer |
| 12 | 20s | Male | White | 227 | 25 | Pfizer |
| 13 | 50s | Male | White | 223 | 26 | Pfizer |
| **N=13 median (IQR) or %** | 45 (32-51) | 92% Male | 85% White | 236 (227-241) | 19 (16-22) | 92% Pfizer |

**Table S3**

Reagents used in the B cell staining panel

| Antibody | Manufacturer | Clone | Catalog |
|---|---|---|---|
| CD3 BV510 | BD Biosciences | HIT3a | 564713 |
| CD14 BV510 | BD Biosciences | MφP9 | 563079 |
| CD56 BV510 | BD Biosciences | NCAM16.2 | 563041 |
| CD19 BUV395 | BD Biosciences | SJ25-C1 | 563549 |
| CD20 BUV737 | BD Biosciences | 2H7 | 564432 |
| CD21 PE-Cy7 | BD Biosciences | B-ly4 | 561374 |
| CD27 BV605 | BioLegend | O323 | 302830 |
| CD38 BB700 | BD Biosciences | HIT2 | 566445 |
| IgA VioBlue | Miltenyi Biotec | IS11-8E10 | 130-114-005 |
| IgD BV650 | BD Biosciences | IA6-2 | 740594 |
| IgG BV786 | BD Biosciences | G18-145 | 564230 |
| IgM PE/Dazzle 594 | BioLegend | MHM-88 | 314530 |
| R-Phycoerythrin Streptavidin | Invitrogen | N/A | S21388 |
| AlexaFluor 488 Streptavidin | Invitrogen | N/A | S32354 |
| AlexaFluor 647 Streptavidin | Invitrogen | N/A | S32357 |
| LIVE/DEAD Fixable Aqua Stain | Invitrogen | N/A | L34957 |
| CD19 Biotin (used as a control) | BD Biosciences | HIB19 | 555411 |

APC, allophycocyanin; BB, brilliant blue; BUV, brilliant ultraviolet; BV, brilliant violet; Cy, cyanine; FITC, fluorescein isothiocyanate; PE R-phycoerythrin; UViD, Live/Dead fixable ultraviolet dead cell stain

**Table S4**

T cell intracellular cytokine staining flow cytometry panel

| Specificity | Fluorochrome | Clone | Vendor | Catalogue |
|---|---|---|---|---|
| Perforin | FITC | B-D48 | BioLegend | 353310 |
| IL-5 | BB630 | TRFK5 | BD | Custom |
| IL-13 | BB630 | JES10-5A2 | BD | Custom |
| Ki67 | BB660 | B56 | BD | Custom |
| IL-4 | BB700 | MP4-25D2 | BD | Custom |
| CRTh2 | PE | BM16 | BioLegend | 350106 |
| CD32 | PE-Dazzle594 | FUN-2 | BioLegend | 303218 |
| CXCR3 (CD183) | PE-Cy5 | 1C6/CXCR3 | BD | 551128 |
| FOXP3 | PE-Cy5.5 | PCH101 | Invitrogen | 35-4776-42 |
| IL-17a | PE-Cy7 | BL168 | BioLegend | 512315 |
| IL-2 | APC | MQ1-17H12 | BioLegend | 500310 |
| Granzyme B | Alexa 700 | GB11 | BD | 560213 |
| CD3 | APC-Fire750 | UCHT1 | BioLegend | 300470 |
| TNF | BUV395 | MAb11 | BD | 563996 |
| Viability | UViD | N/A | Invitrogen | 65-0863 |
| CD45RA | BUV496 | HI100 | BD | 750258 |
| CD19 | BUV563 | SJ25C1 | BD | 612916 |
| CD14 | BUV661 | MΦP9 | BD | 741684 |
| CD154 | BUV737 | TRAP1 | BD | 748983 |
| CD8 | BUV805 | SK1 | BD | 612889 |
| IFNγ | V450 | B27 | BD | 560371 |
| CD4 | BV480 | SK3 | BD | 566104 |
| CD16 | BV570 | 3G8 | BioLegend | 302036 |
| CCR7 | BV605 | G034H7 | BioLegend | 353224 |
| CD25 | BV650 | M-A251 | BD | 563719 |
| CD64 | BV711 | 10.1 | BioLegend | 305042 |
| CD56 | BV750 | 5.1H11 | BioLegend | 362556 |
| CCR6 (CD196) | BV786 | 11A9 | BD | 563704 |

APC, allophycocyanin; BB, brilliant blue; BUV, brilliant ultraviolet; BV, brilliant violet; Cy, cyanine; FITC, fluorescein isothiocyanate; PE R-phycoerythrin; UViD, Live/Dead fixable ultraviolet dead cell stain
